# The interactive effects of ambient air pollutants-meteorological factors on confirmed cases of COVID-19 in 120 Chinese cities

**DOI:** 10.1101/2020.05.27.20111542

**Authors:** Jianli Zhou, Linyuan Qin, Xiaojing Meng, Nan Liu

**Affiliations:** Department of Occupational Health and Occupational Medicine, Guangdong Provincial Key Laboratory of Tropical Disease Research, School of Public Health, Southern Medical University, Guangzhou, 510515, P. R. China; Department of Epidemiology and Statistics, School of Public Health, Guilin Medical University, Guilin, 541001, P. R. China; Pinghu Hospital, Health Science Center, Shenzhen University, Shenzhen, 518116, P. R. China; Institute of Public Health, School of Nursing, Henan University, Kaifeng, 475004, P. R. China; College of Public Health, Zhengzhou University, Zhengzhou, 540001, P. R. China

**Keywords:** air pollutants, COVID-19, interactive effect, meteorological parameters, migration scale index, negative binomial regression

## Abstract

Emerging evidence has confirmed meteorological factors and air pollutants affect novel coronavirus disease 2019 (COVID-19). However, no studies to date have considered the impact of interactions between meteorological factors and air pollutants on COVID-19 transmission. This study explores the association between ambient air pollutants, meteorological factors and their interaction on confirmed case counts of COVID-19 in 120 Chinese cities. We modeled total confirmed cases of COVID-19 with meteorological factors, air pollutants and their interactions. To account for potential migration effects, we included the migration scale index (MSI) from Wuhan to each of the 120 cities included in the model, using data from 15 January to 18 March 2020. As an important confounding factor, MSI was considered in a negative binomial regression analysis. Positive associations were found between the number of confirmed cases of COVID-19 and carbon monoxide, aerodynamic particulate matter with aerodynamic diameter ≤2.5 µm, relative humidity and ozone, with and without MSI-adjustment. Negative associations were also found for sulfur dioxide and wind velocity both with and without controlling for population migration. In addition, air pollutants and meteorological factors had interactive effects on COVID-19 after controlling for MSI. In conclusion, air pollutants, meteorological factors and their interactions all affect COVID-19 cases.

## 1. Introduction

In Dec. 2019, a novel coronavirus disease (COVID-19) caused by severe acute respiratory syndrome coronavirus 2 (SARS-CoV-2) was reported in Wuhan City, P. R. China (Wu et al. 2020, Zhou et al. 2020). The COVID-19 outbreak rapidly spread across China and to many other countries (Holshue et al. 2020, Huang et al. 2017, Lai et al. 2020, Wang et al. 2020a). Due to its transmissibility and increasing spread, COVID-19 was officially declared a pandemic by the World Health Organization (WHO) on 11 March 2020 (https://www.who.int/emergencies/diseases/novel-coronavirus-2019). By 17 August 2020, COVID-19 had become prevalent in 188 countries, with a total of 21,869,433 confirmed cases worldwide (https://www.who.int/emergencies/diseases/novel-coronavirus-2019/situation-reports/); and 89906 confirmed cases in China (http://www.nhc.gov.cn/xcs/xxgzbd/gzbd_index.shtml). This has created a worldwide public health crisis.

Air pollutants can significantly affect health and the WHO indicates that 4.6 million individuals die annually from diseases and illnesses directly related to poor air quality (Tobias &Molina 2020). In particular, increasing numbers of studies suggest a relationship between air pollution and infectious diseases. While previous studies report no relationship between air pollution and SARS outbreaks (Cai et al. 2007, Tobias &Molina 2020, Yusuf et al. 2007), a positive association was observed between air pollutants (sulfur dioxide, nitrogen dioxide, carbon monoxide and ground-level ozone) and SARS fatality rates (Cui et al. 2003). Another study in Brisbane suggested that high concentrations of ozone (O_3_) and particulate matter <10 microns (PM_10_) were associated with increased pediatric influenza cases (Xu et al. 2013).

Meteorological factors are another independent factor that can affect the spread of infectious diseases (Cai et al. 2007, Xie &Zhu 2020). Although the association between viruses leading to respiratory disease (e.g., SARS, viral influenza and SARS-CoV-2) and meteorological factors has been explored (Yusuf et al. 2007), results of these studies remain controversial. For example, Tan et al. found a sharp change in ambient temperature was correlated with increased risk of SARS, (Tan et al. 2005), while Yao et al. found ambient temperature had no significant effect on the transmissibility of SARS-CoV-2 (Yao et al. 2020b). Other studies indicate that an increase in average temperature contributes to an increase in average relative risk (RR) but a decrease in daily confirmed case counts of SARS (Cai et al. 2007). These inconsistent results may reflect an interaction occurring between pollutants and meteorological effects on the transmission of these diseases.

The trends of the SARS outbreak may be influenced by air pollutants and meteorological factors that change host susceptibility and survival time of SARS-CoV in vitro (Cai et al. 2007). COVID-19 infections can cause serious respiratory disease. Common symptoms at the beginning of infection include fever, cough, muscle aches or fatigue, etc., similar to SARS-CoV (Liu et al. 2020b, Siordia 2020), and an effect of air pollutants and meteorological factors, similar to that seen in SARS, on confirmed COVID-19 cases has also observed (Jiang et al. 2020, Liu et al. 2020a, Oliveiros et al. 2020, Wang et al. 2020b, Zhu et al. 2020); However, the interactive effects of air pollutants and meteorological factors has not yet been considered. Given the human to human transmission of COVID-19, population migration is also likely to be an important confounding factor when evaluating the effects of meteorological factors and air pollutants on COVID-19, and it has been established that COVID19 cases are almost linearly correlated with the number of people traveled from Wuhan to the destination city (Hu Jianxiong 2020). In this study, we investigate the association between ambient air pollutants, meteorological factors, and their interactive effects on the number of COVID-19 confirmed cases while also controlling for population migration effects, to extend and further our understanding of the relationship between transmission of this disease and environmental factors.

## 2. Methods

### 2.1. Data collection

We observed the numbers of the confirmed cases in relation to air pollutants, meteorological parameters and the migration scale index (MSI) in 120 Chinese cities covering the majority of mainland China. The MSI reflects the scale of urban migration into or out of the assigned population, using a horizontal comparison among cities to reflect the trend of population migration between the city pairs. All cities included had no less than five cases from 15 January to 18 March 2020 inclusive. Wuhan was locked down at 10 am on 23 Jan, so we consider only people that traveled from Wuhan to the destination city between 17 and 23 Jan. 2020. All data were taken from the official website of Baidu Migration (https://qianxi.baidu.com/2020/). To better fit the model, we do not include the data of Wuhan City since its large number of the confirmed case counts skew the data and thus may reduce the robustness of the model.

Daily confirmed case counts and daily air pollutants were collected from the official website of Harvard University (https://dataverse.harvard.edu). Meteorological indexes, including average temperature (AT), diurnal temperature range (DTR), relative humidity (RH), wind velocity (WV), air pressure (AP), precipitation (PRE), and hours of sunshine (HS), were obtained from China’s Meteorological Data Sharing Service System (http://data.cma.cn/site/index.html). Air pollutant data included particulate matter with aerodynamic diameter ≤2.5 µm (PM_2.5_), nitrogen dioxide (NO_2_), sulfur dioxide (SO_2_), carbon monoxide (CO), and ozone (O_3_). Pollutant concentrations and meteorology parameters were available as daily means in each city; we calculated a single mean for the 63 days of our date range for use in the model.

### 2.2 Statistical analysis

Descriptive analysis was performed on all data to explore the characteristics of total confirmed case counts, average concentration of O_3_, NO_2_, SO_2_, CO, PM_2.5_, AT, DTR, RH, WV, AP, PRE, HS and MSI. Meanwhile, to account for the lag effect and the latent period of COVID-19, we evaluated associations with confirmed case counts using lags of 0, 3, 7 and 14 d for air pollutants and meteorological factors (Liu et al. 2020a). Lags were incorporated into parameters by shifting the date range of the appropriate number of days. For example, for lag 3, mean parameter values were calculated using daily means from 15 Jan to 15 March for use in the model; confirmed case counts were always for the date range 18 Jan to 18 March. Spearman correlation analysis was employed to explore the correlation between meteorological factors and air pollutants, and among air pollutants, meteorological factors and confirmed case counts. We used generalized linear models with a negative binomial error distribution to analyze associations between air pollutant factors (O_3_, NO_2_, SO_2_, CO and PM_2.5_), meteorological factors (AT, DTR, RH, WV, AP, PRE and HS), MSI and the total counts of confirmed case since the count data was expected to be clustered and thus over-dispersed. First, we established simple negative binomial regression models (including only one independent variable) to analyze each air pollutant, meteorological factor, and MSI effects on confirmed case counts. We then conducted multiple negative binomial regression models (including multiple independent variables), with and without MSI, that included all above-mentioned air pollutants and meteorological factors since the independent variables are not strongly correlated (all |r| <0.9). The general negative binomial regression equation is

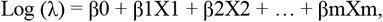

where *λ* represents the risk of being a confirmed case counts, *X*1, *X*2, …, *Xm* are the factors that affect *λ*; and β1, β2, …, β*m* are the partial coefficients of *X*1, *X*2, and *Xm*, respectively. To explore the interaction effects, we included *X*i\**Xj* as an interaction term in the model, where *Xi* and *Xj* represent an air pollutant and a meteorological factor, respectively. A stratified analysis was done to consider the effect of air pollutants on confirmed COVID-19 cases under different levels of meteorological factors. When a significant interaction effect was present in the negative binomial model, the meteorological factor was stratified into two groups using the median as the dividing point, and a negative binomial model containing all pollutants and remaining meteorological factors was fitted to analyze the effect of air pollutants on the confirmed number of COVID-19 cases was done for each group.

All statistical analyses were performed using SPSS version 22.0 (SPSS Inc., Chicago, IL, USA) and R Core Team (2020). R: A language and environment for statistical computing. R Foundation for Statistical Computing, Vienna, Austria. URL http://www.R-project.org/ with a two-sided P-value < 0.05 considered as statistically significant.

## 3. Results

### 3.1 Study area

Figure 1(a) displays the location of the 120 selected cities in mainland China and the total confirmed cases in each city collected from 15 January 18 March 2020. The latitude and longitude of these 120 cities range from 20°00′ to 47°23′ N and from 99° 11′ to 130°55′ E, respectively. Most cases were concentrated in the central and eastern parts of mainland China, with fewer cases in the northwest. The detailed distribution of confirmed COVID-19 cases in the 120 cities is given in Table S1. Hubei province contained the majority of confirmed COVID-19 cases; excluding Wuhan City, Xiangfan, Shiyan, Yichang, Jingzhou, Suizhou, and Xiaogan were the cities with the highest number of cases (>500 per city).

**Figure 1.**
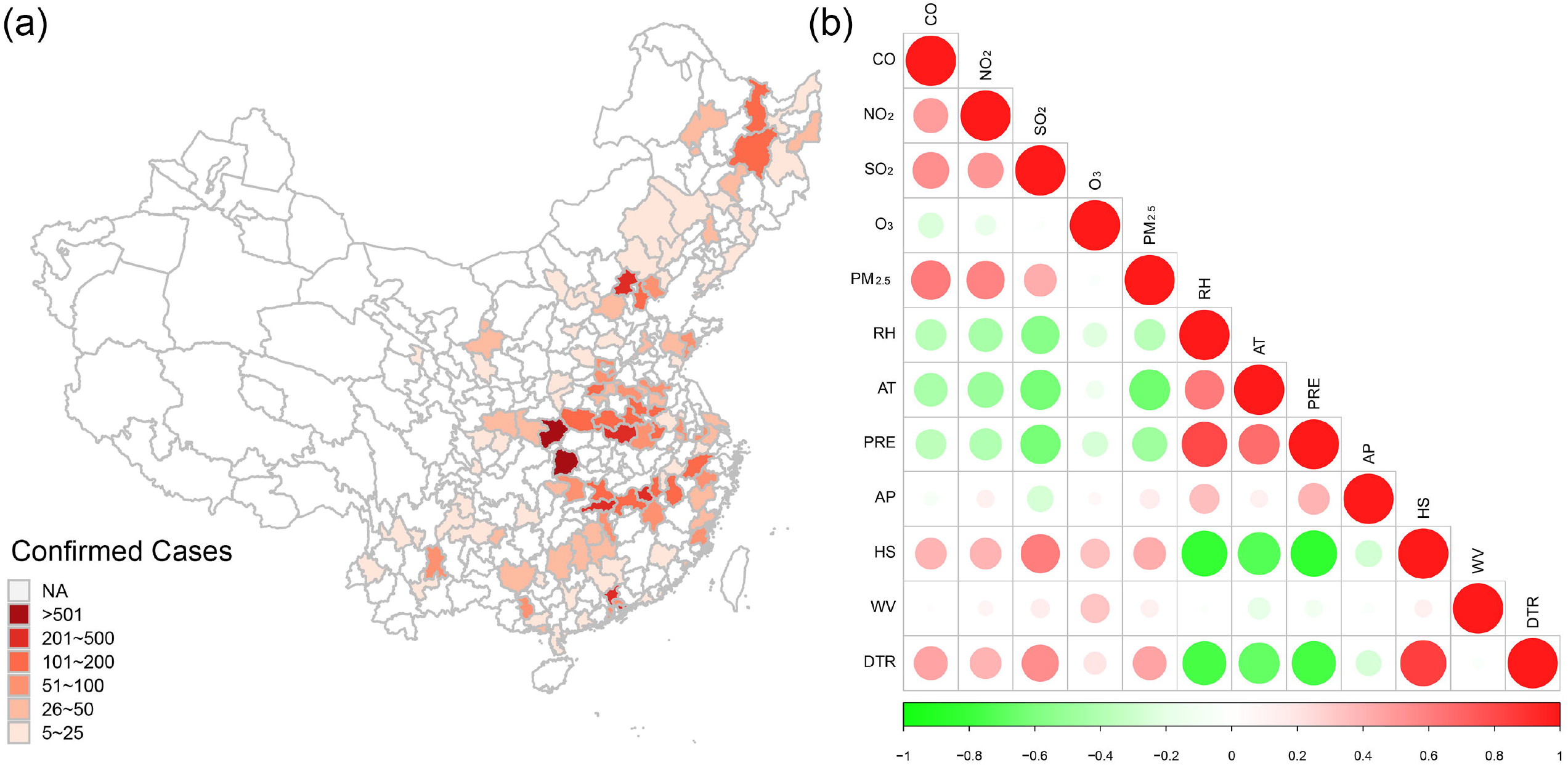
Locations of 120 cities and total confirmed cases in each city on 18 Mar. 2020, and correlation analysis of meteorological factors and air pollutants. NA: Cities not included in this study.

### 3.2 Description of confirmed COVID-19 cases, meteorological factors and air pollutants

Descriptive statistics (maximum and minimum values, median, and interquartile range) for the number of confirmed cases, air pollutant measures, and the meteorological parameters of the 120 cities are presented in Table 1. Among these cities, during our study period (15 January to 18 March 2020), the median daily concentrations of CO, NO_2_, SO_2_, O_3_ and PM_2.5_, were 0.82 mg/m^3^, 20.89 μg/m^3^, 8.23 μg/m^3^, 56.38 μg/m^3^, and 43.58 μg/m^3^, respectively. For meteorological parameters, ranges of RH, AT, PRE, AP, HS, WV, DTR were 35.42-85.50%, −12.48 to 21.29 °C, 0.00-6.82 mm, 758.41-1024.43 hPa, 1.60-9.20 h, 1.00-5.05 m/s, and 5.13 to 18.37 °C, respectively. MSI ranged from 0.01 to 5.13.

**Table 1.**
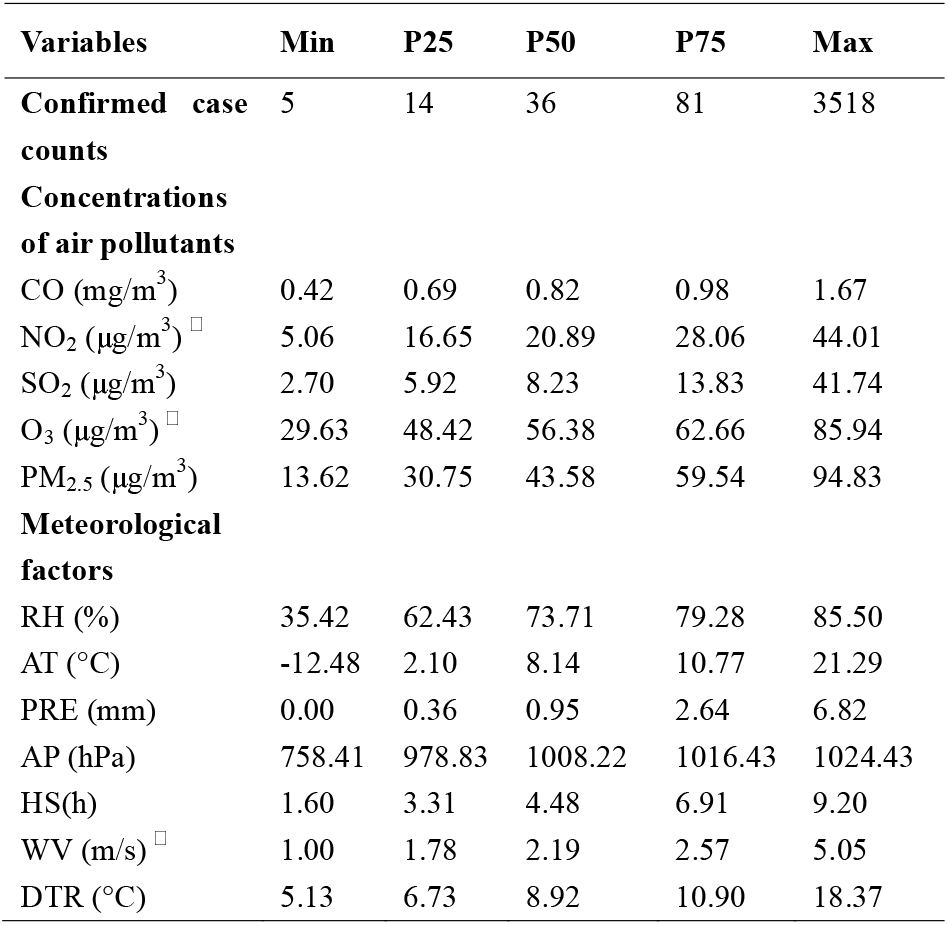
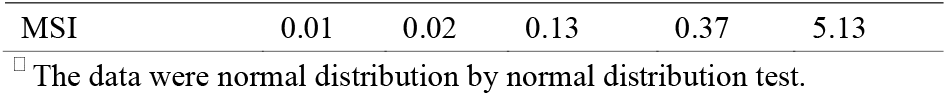
Summary of total confirmed COVID-19 cases, meteorological data and air pollutants of 120 Chinese cities

### 3.3 Correlation analysis of air pollutants and meteorological factors

Spearman correlation coefficients (r_s_) between concentrations of air pollutants and meteorological factors are shown in Figure 1(b). Circle size indicates strength of relationship between two variables. All correlations are less than |0.9|. Therefore, all the variables could be integrated into the multiple negative binomial regression model.

### 3.4 Effects of air pollutants and meteorological parameters on confirmed case counts

The relationship between pollutants, meteorological parameters and confirmed case counts for the lag of 14 d are presented in a scatter plot in Figure 2. Positive associations were present between the confirmed case counts and the pollutant measure PM_2.5_ (*P* = 0.024), and the meteorological measures RH (*P* = 0.001), PRE (*P* = 0.015) and AP (*P* <0.001). Negative associations occurred between the confirmed case counts and the pollutant SO_2_ (*P* <0.001) and the meteorological variable HS (*P* = 0.018). Correlations between confirmed case counts and all other variables were not significant.

**Figure 2.**
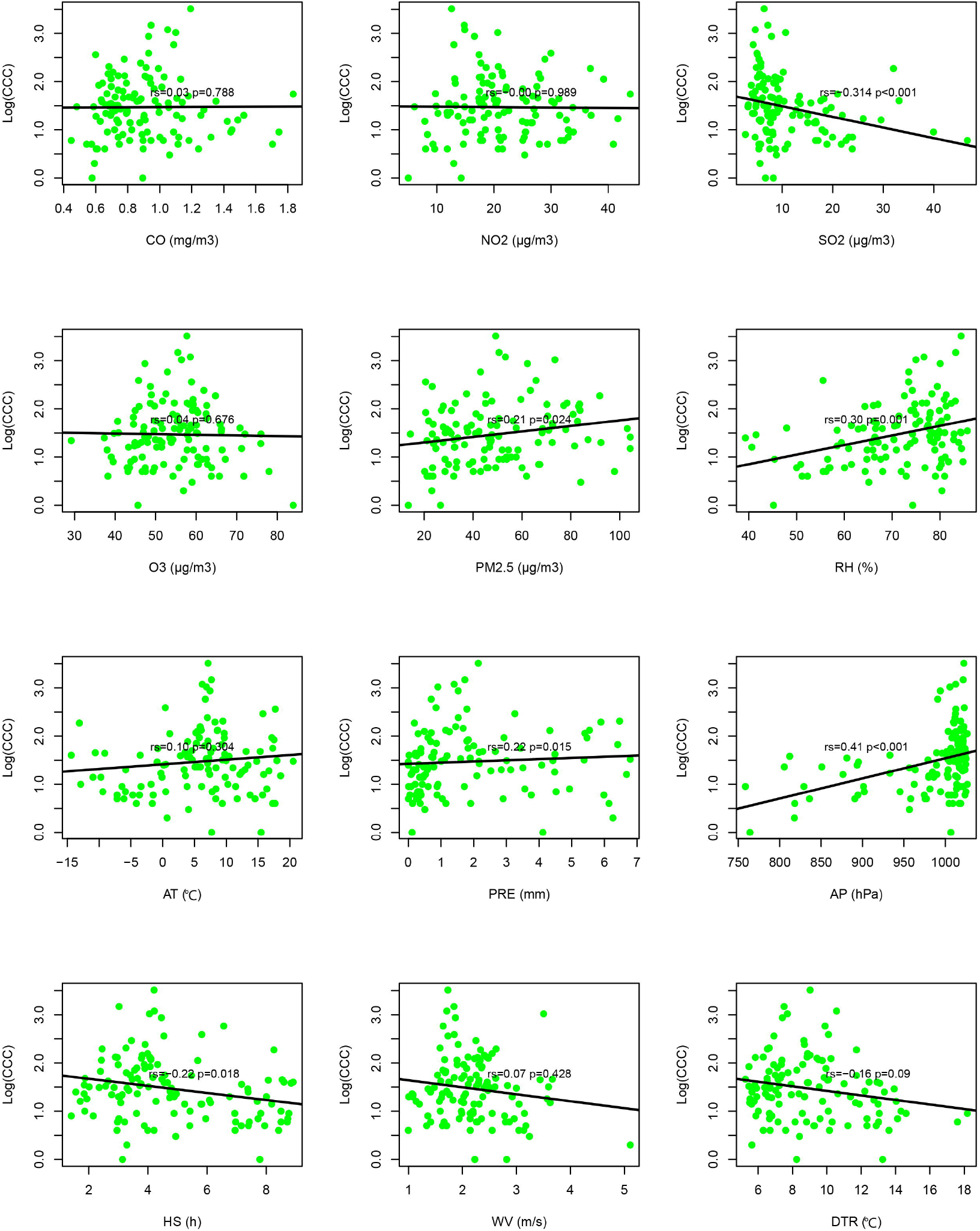
Correlation analysis of confirmed case counts with air pollution and meteorological parameters. Log (CCC) = log (confirmed case counts)

### 3.5 Negative binomial regression model analysis

Simple negative binomial regression results are shown in Figure S1(a-d). CO, NO_2_, SO_2_, O_3_, PM_2.5_, RH, AT, AP, HS, WV and MSI were significantly associated with confirmed cases counts (all *P* <0.05). PRE had no effect on confirmed case counts for lags of 0, 3, 7 and 14 d, while DTR was significantly associated with confirmed cases counts at lags of 7 and 14 d. The multiple negative binomial regression model results (without adjusting for MSI) are shown in Figure 3(a1-d1). CO, PM_2.5_ and RH were positively associated with confirmed case counts, while SO_2_ was negatively associated with confirmed case counts at lags 0, 3, 7, and 14 d. However, O_3_ has a positive effect on confirmed case counts at lag 0 and WV has a positive effect on confirmed case counts at lag 14 d. After adjusting for migration using MSI, CO, PM_2.5_, RH, O_3_, SO_2_, WV were also statistically significant, as shown in Figure 3(a2-d2).

**Figure 3.**
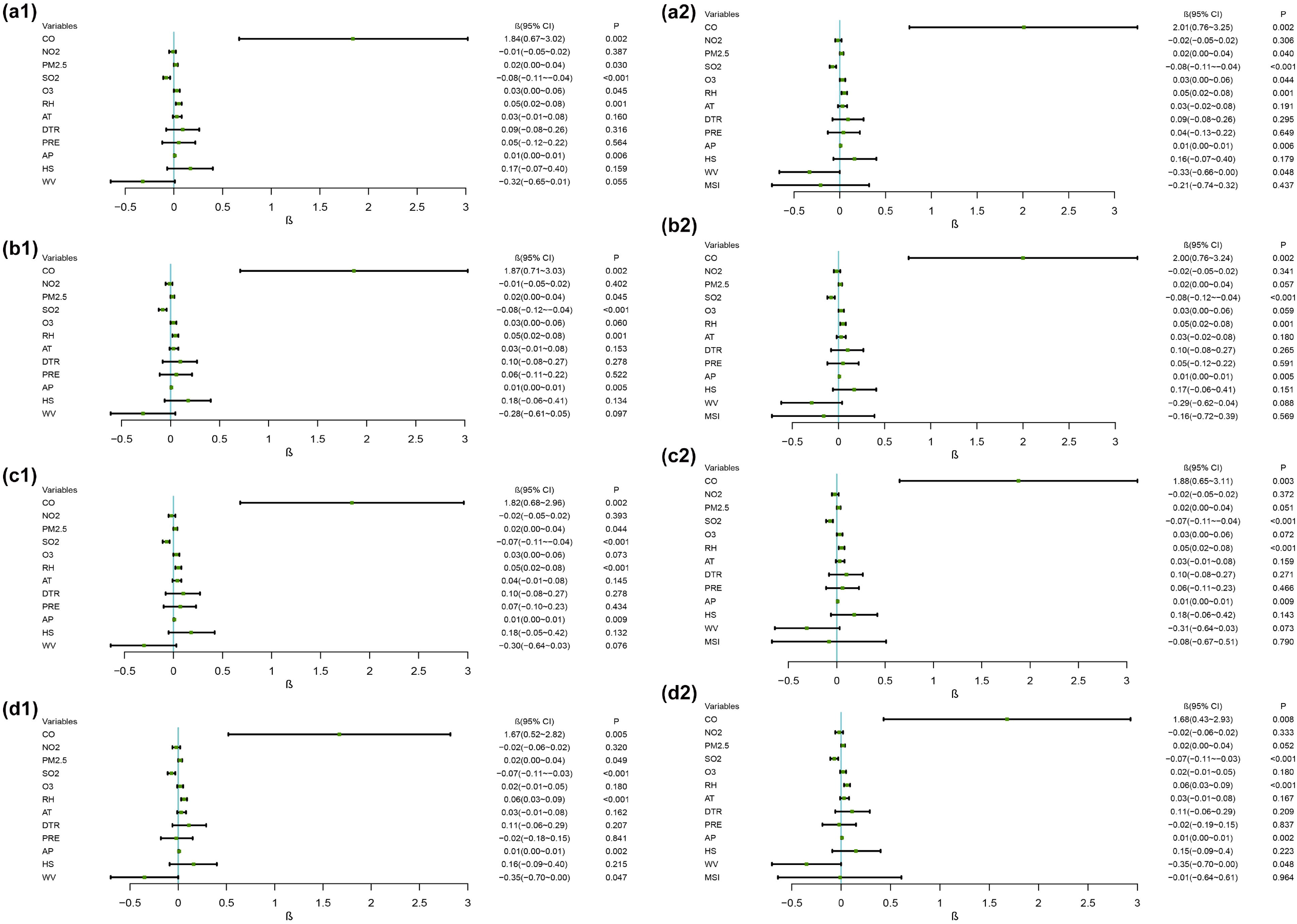
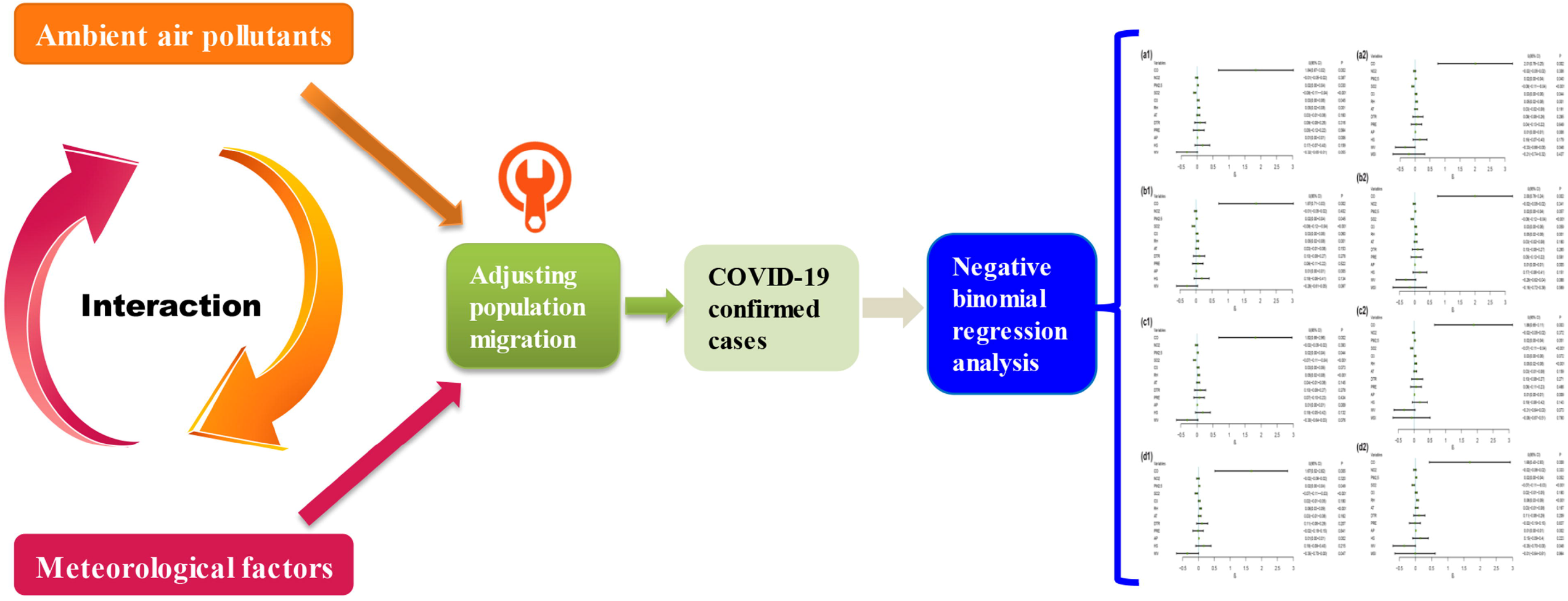
The relationship between air pollutants, meteorological factors and confirmed case counts in a multiple negative binomial regression model with and without adjusting for migrations with MSI.

### 3.6 Interaction effects between air pollutants and meteorological factors

Significant interactive effects (*P* <0.05) of ambient air pollutants and meteorological factors on COVID-19 confirmed cases after adjusting with MSI are shown in Table 2. CO interacted with two meteorological factors, HS and DTR, and SO_2_ interacted with AP to negatively affect the total number of COVID-19 confirmed cases; while CO and RH, and PM_2.5_ and PRE interacted to positively affect total case count.

**Table 2.**
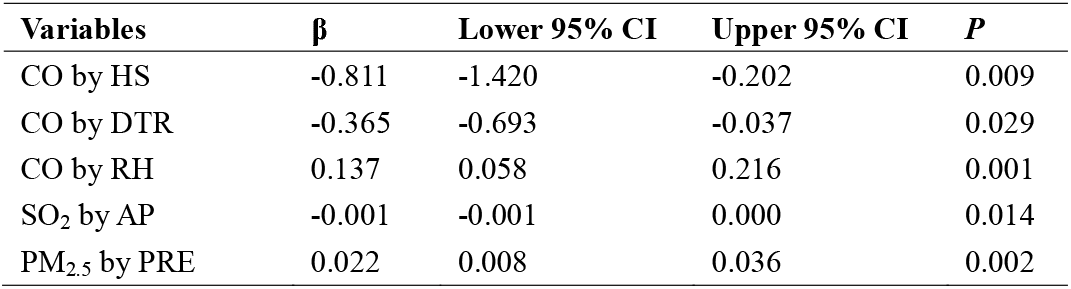
The significant interaction effects of ambient air pollutants and meteorological factors on COVID-19 confirmed cases after adjusting MSI (*P*<0.05)

The adjusted interaction effects of CO with HS, DTR, RH, of SO_2_ with AP, and of PM_2.5_ with PRE are listed in Table 3. The remaining adjusted air pollutants, meteorological factors and MSI and under the levels of HS values below 50^th^ percentile and HS and above 50^th^ percentile, had no significant effects on COVID-19 case counts by CO. At the low DTR level (<50^th^ percentile), an increase of CO by 1 mg/m^3^ is expected to increase the log of risk of being confirmed COVID-19 cases by a factor of >5 (β = 5.763, *P* = 0.001), while the same increase in CO at the high DTR level (≥ 50^th^ percentile) increases the riskby a factor of <1 (β = 0.931, *P* = 0.035). CO has no significant effect on confirmed case counts at low RH level, but at a high RH level, a 1 mg/m^3^ increase of CO significantly raises the risk of being confirmed case (β = 6.045, *P* <0.001). An increase of SO_2_ was associated with lower risk of confirmed case counts (β = −0.243, *P* <0.001) at high level AP, but was not significantly associated with confirmed case counts at low level AP. PM_2.5_ variation had no significant effect for either low or high PRE levels. These results indicate that the effects of CO and SO_2_ vary depending on the level of meteorological factors.

**Table 3.** Stratified analysis of ambient air pollutants and meteorological factors interaction effects on COVID-19 confirmed cases

| Air pollutant | Meteorological factor | $\beta$ | Lower 95% CI | Upper 95% CI | *P* |
| --- | --- | --- | --- | --- | --- |
| CO | HS<P50 | 3.321 | -0.230 | 6.873 | 0.067 |
|  | HS≥P50 | 1.115 | -0.247 | 2.478 | 0.108 |
| CO | DTR<P50 | 5.763 | 2.402 | 9.124 | 0.001 |
|  | DTR≥P50 | 0.931 | 0.066 | 1.796 | 0.035 |
| CO | RH<P50 | 0.930 | -0.574 | 2.434 | 0.226 |
|  | RH≥P50 | 6.045 | 3.247 | 8.844 | <0.001 |
| SO<sub>2</sub> | AP<P50 | -0.006 | -0.052 | 0.041 | 0.812 |
|  | AP≥P50 | -0.243 | -0.307 | -0.178 | <0.001 |
| PM<sub>2.5</sub> | PRE<P50 | 0.004 | -0.019 | 0.027 | 0.727 |
|  | PRE≥P50 | 0.049 | -0.009 | 0.107 | 0.100 |

## 4. Discussion

In this work, we explored the role of air pollutants, meteorological factors and their interactive effects on confirmed case counts in 120 Chinese cities. We focused on these cities as their meteorological data was available (this is not true for all cities in China). Our results indicate that air pollutants, meteorological factors and their interaction are associated with COVID-19 confirmed case counts, with and without adjusting using MSI.

Previous studies indicate that meteorological factors and air pollutants influence prevalence of infectious diseases (e.g., influenza, SARS, MERS, COVID-19 (Cai et al. 2007, Liu et al. 2020a, Prata et al. 2020, Tan et al. 2005, van Doremalen et al. 2013, Yusuf et al. 2007)). An ecological analysis carried out in China demonstrated that air pollutants increase SARS case fatalities (Cui et al. 2003). As a major component of air pollution, particulate matter is associated with a variety of cardiorespiratory diseases, including acute lower respiratory tract infections. PM_2.5_ has adverse effects on human health and is associated with many diseases, especially respiratory disease (Horne et al. 2018). PM_2.5_ may damage bronchial immunity and epithelial cell integrity (Jiang et al. 2020), which, in turn, will reduce the body’s ability to fight off viruses, making affected individuals more vulnerable to respiratory disease. Exposure to higher concentrations of PM_2.5_ results in greater use of healthcare resources for acute lower respiratory infection (Horne et al. 2018). Jiang et al. (Jiang et al. 2020), Suhaimi et al. (Suhaimi et al. 2020) and Zhu et al. (Zhu et al. 2020) studies suggested that PM_2.5_ was positively associated with confirmed COVID-19 cases, which were consistent with our results (Figure 3). Cai et al. (2007) indicated that no correlation exists between air pollution (including particulate matter, SO_2_, NO_2_, O_3_, and CO) and SARS outbreaks (Cai et al. 2007). However, our results are consistent with Zhu et al. (Zhu et al. 2020), who found that increases in CO were positively associated with the number of confirmed COVID-19 cases, while SO_2_ was negatively correlated with COVID-19 cases (Figure 3). The difference is that migration was taken into account in our analysis and it was not in the analysis by Zhu et al. (Zhu et al. 2020). Hu’s (Hu Jianxiong 2020) study indicated that the cumulative number of reported COVID-19 cases in each province was positively correlated with the population index of Wuhan moving out to each province, and the correlation coefficient is 0.84, so we believe that it is necessary to control the population migration.

Kesic et al. suggests that exposure to O_3_ might disrupt the protease/anti-protease balance of human airways, contributing to an increased influenza A infection rate (Kesic et al. 2012). We similarly found that exposure to PM_2.5_, CO and O_3_ increased likelihood of being infected by COVID-19 (Figure 3). Understanding the relationship between air pollutant exposure and health provides clues regarding effective protective measures that can reduce the incidence or risk of diseases. Despite the positive relationship between air pollutants (including PM_2.5_, CO, and O_3_) and confirmed COVID-19 cases and the negative relationship between SO_2_ and confirmed COVID-19 cases that we found, the mechanism by which pollutants affect COVID-19, whether via changes in the transmission of the virus or in the immunity of the human body, remains unknown. Further experimental studies are required to explore the molecular regulation mechanisms involved.

Meteorological factors such as AT, DTR, RH, WV, AP, PRE and HS are important factors influencing the survival of microorganisms in the external environment (Anwar et al. 2019, Bezirtzoglou et al. 2011, Casadevall 2020, Chan et al. 2011, Liu et al. 2020a). For decades, experts have issued warnings that climate change may affect the epidemiology of infectious diseases (Casadevall 2020). Global warming, rising sea levels, and climate change will be accompanied by new and unknown diseases (Bezirtzoglou et al. 2011, Casadevall 2020). Meteorological factors may change the host’s behavior, e.g., by affecting time spent indoors or outdoors, affect the host’s defense mechanisms (inhalation of dry and cold air will cause damage to the cleaning function of cilia), or change the infectivity and stability of the virus. Although many studies on meteorological factors and respiratory infectious diseases (including COVID-19) exist, research results remain controversial. While temperature is unlikely to act alone in determining the population’s health, but it can work synergistically with other weather parameters to do so (Huang et al. 2017).

COVID-19 case numbers appear to increase with increasing temperature in the range of −20 to 20 °C (Liu et al. 2020a). Other studies have reported that when AT is below 3 °C, it is positively correlated with confirmed COVID-19 case counts (Xie &Zhu 2020). A Brazilian study suggests a negative linear relationship exists between AT and daily cumulative confirmed cases of COVID-19 in the temperature range of 16.8 to 27.4 °C (Prata et al. 2020), while Auler et al.’s research (also conducted in Brazilian) suggests both higher AT and RH favor COVID-19 transmission.(Auler et al. 2020). The results of studies in some tropical countries emphasize that COVID-19 cases positively correlate with AT (Pani et al. 2020, Suhaimi et al. 2020, Tosepu et al. 2020). Another study demonstrated that in DTR increased in range of from 5 to 15 °C, the counts of confirmed COVID-19 cases decreased, and that a non-linear relationship occurred between COVID-19 cases and absolute humidity (Liu et al. 2020a). A study of 244 cities in China found no significant associations between RH, AT and cumulative incidence rate or R_0_ of COVID-19 (Yao et al. 2020a), which is in contrast to our finding that RH and AT are positively associated with COVID-19 case counts (Figure 3 and S1, respectively). Temperature in our study ranged from −12.48 to 21.29 °C, and our results of AT as positively associated with confirmed case counts is consistent with other studies in tropical countries and China indicating that COVID-19 can survive at higher temperatures, this reveals that COVID-19 can survival under higher temperatures. Experimental research on SARS-CoV-2 confirmed that this virus is very stable at 4 °C, but when the temperature increases to 70 °C, inactivation time for the virus is shortened to 5 min (Chin et al. 2020).

Hu et al. reported that low WV might increase the risk of mumps (Hu et al. 2018). Our multiple negative binomial regression models [lag 14 d in Figure 3(d1)] suggest that WV is negatively associated with COVID-19 confirmed case counts, consistent with Hu et al (Hu et al. 2018). Wind can affect the duration of droplets suspended in air. In indoor environments, high wind speed means that rooms are well-ventilated and in outdoor environments, high wind speed enhances dilution and removal of droplets, both of which could shorten droplet suspension time in the air (Cai et al. 2007).

Regarding rainfall, our study showed PRE is positively related to confirmed case counts (Figure 2); however, this effect was not statistically significant in simple and multiple negative binomial regression analysis (Figure S1 and 3). Some epidemiological data generally support a relationship between influenza virus infection and PRE (Pica &Bouvier 2012). Agrawal et al. found a significant association between rainfall and influenza virus infection in India, and fewer reports of infection in the dry season (Agrawal et al. 2009). The association between PRE and respiratory syncytial virus infection showed similar results in our research and in other studies (Shek &Lee 2003).

To the best of our knowledge, interactive effects of air pollutants and meteorological factors have not previously been identified as affecting COVID-19 cases but have been identified in noninfectious diseases (Guo et al. 2019, Huang et al. 2017) and cause-specific diseases (Chen et al. 2019). In this study, we observed interactive effects on the total number of COVID-19 confirmed cases, especially for CO, which showed interactions with three meteorological factors (HS, DTR and RH; Table 2). The degree of increase in number of COVID-19 confirmed cases with CO depends on the level of certain meteorological factors. For example, effect of CO on confirmed cases is 5.763 (β *=* 5.763, *P*= 0.001) at low DTR level (< 50th percentile), but it is 0.931 (β*=* 0.931, *P*= 0.035) at high DTR level (Table 3) and the interactive effect was negative (β *=* −0.365, *P* = 0.029, Table 2), creating an antagonistic effect between these two factors. Although the interactive effect of CO and HS was statistically significant (β *=* −0.811, *P* = 0.009, Table 2), the stratified analysis did not find differences between the two categories of HS (Table 3), and thus the biological meaning of the interaction is unclear. Interactive effects of PM_2.5_ with PRE and SO_2_ with AP were also statistically significant. Our findings suggest that environmental pollutants do not influence COVID-19 spread alone, but that they play important roles in combination with meteorological variables. The exact mechanisms of these interactive effects requires for further study.

We have comprehensively evaluated the effect of environmental pollution, meteorological factors and their interaction on the number of confirmed COVID-19 cases with and without controlling for MSI, to increase understanding of these risk factors on the transmission of COVID-19. The limitation of our research is that we only analyzed data from 120 cities in China. Further studies are required in more countries and regions to clarify the effect of environmental air pollutants on COVID-19 infection rates worldwide.

## 5. Conclusion

Our work verifies that air pollutants, meteorological factors and their interaction affect the number of confirmed cases of COVID-19 even after controlling for migration. Our study suggests that increased RH, CO and PM_2.5_ concentrations are associated with increased numbers of confirmed COVID-19 cases. In addition, pollutant-meteorological factor interaction effects influenced COVID-19 infection levels. Our research provides a theoretical basis for formulating public health policies for the prevention and control of COVID-19 that take into consideration the effects and relationship of air pollutants and meteorological factors.

## Data Availability

The [COVID-19 confirmed cases counts and air pollutants] data used to support the findings of this study were freely available from https://dataverse.harvard.edu.
The [meteorological factors] data used to support the findings of this study were freely available from http://data.cma.cn/site/index.html if you registered users with real names in education and scientific research.

## Acknowledgments

This work was supported by National Natural Science Foundation of China (Nos. 81872584 and 81273078), National 863 Young Scientist Program (No. 2015AA020940), Science and Technology Program of Guangzhou (No. 201704020056), and Scientific Research Project for University of Education Bureau of Guangzhou (No. 201831841).

## Conflict of interest

The authors declare that they have no conflict of interest.

## Author contributions

Jianli Zhou: conceptualization, methodology, data curation, writing-original draft preparation. Linyuan Qin: methodology, visualization, investigation, writing-original draft preparation. Xiaojing Meng: writing-reviewing and editing. Nan Liu: supervision, validation, writing-reviewing and editing.

## Appendix A. Supplementary materials

Supplementary materials to this article can be found online at…

## Notes

### Competing Interest Statement

The authors have declared no competing interest.

### Author Declarations

Data is obtained from the official website, therefore, there is no need to have ethical review.

