## Supplementary Materials for "The interactive effects of ambient air pollutants-meteorological factors on confirmed cases of COVID-19 in 120 Chinese cities"

Electronic Supplementary Material

on *Environmental Science and Pollution Research* publication entitled

Jianli Zhou ^a^, Linyuan Qin ^b^, Xiaojing Meng ^a*^, Nan Liu ^a, c, d, e*^
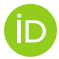

^a^ Department of Occupational Health and Occupational Medicine, Guangdong Provincial Key Laboratory of Tropical Disease Research, School of Public Health, Southern Medical University, Guangzhou, 510515, P. R. China.

^b^ Department of Epidemiology and Statistics, School of Public Health, Guilin Medical University, Guilin, 541001, P. R. China.

^c^ Pinghu Hospital, Health Science Center, Shenzhen University, Shenzhen, 518116, P. R. China.

^d^ Institute of Public Health, School of Nursing, Henan University, Kaifeng, 475004, P. R. China.

^e^ College of Public Health, Zhengzhou University, Zhengzhou, 540001, P. R. China.

*Corresponding authors:

**Xiaojing Meng**

**Nan Liu**

orcid.org/0000-0002-8895-3169

**Supporting Table**

Table S1. Distribution of confirmed COVID-19 cases in 120 cities of Chinese mainland

| **Confirmed case number** | **Number of cities** | **Belong to province** |
| --- | --- | --- |
| >501 | 6 | Xiangfan, Shiyan, Yichang, Jingzhou, Suizhou, Xiaogan (Hubei Province) |
| 201-500 | 6 | Beijing  Xinyang (Henan Province)  Nanchang (Jiangxi Province)  Changsha (Hunan Province)  Guangzhou and Shenzhen (Guangdong Province) |
| 101-200 | 13 | Tianjin  Hangzhou (Zhejiang Province)  Yueyang (Hunan Province)  Haerbin (Heilongjiang Province)  Shangrao and Yichun (Jiangxi Province)  Hefei, Bengbu, Bozhou and Fuyang (Anhui Province)  Zhengzhou, Nanyang, Zhumadian (Henan Province) |
| 51-100 | 18 | Changde and Tangshan (Hebei Province)  Qingdao (Shandong Province)  Liuan, (Anhui Province)  Fuzhou (Fujian Province)  Kunming (Yunnan Province)  Jinhua (Zhejiang Province)  Nanning (Guangxi Province)  Zhuzhou (Hunan Province)  Anyang, Shangqiu and Xinxiang (Henan Province)  Nanjing, Wuxi and Xuzhou (Jiangsu Province)  Dongguan, Zhongshan, Zhuhai (Guangdong Province) |
| 26-50 | 29 | Changchun (Jilin Province)  Shenyang (Liaoning Province)  Baoding (Hebei Province)  Lishui (Zhejiang Province)  Chenzhou (Sichuan Province)  Guiyang (Guizhou Province)  Haikou (Hainan Province)  Jixi and Qiqihaer (Heilongjiang Province)  Ankang and Hanzhong (Shanxi Province)  Jinan and Weifang (Shandong Province)  Kaifeng and Xuchang (Henan Province)  Yinchuan and Wuzhong (Ningxia Province)  Maanshan and Tongling (Anhui Province)  Nantong and Suzhou (Jiangsu Province)  Ningde and Xiamen (Fujian Province)  Hengyang, Yongzhou and Zhangjiajie (Hunan Province) Beihai, Guilin and Hechi (Guangxi Province) |
| 5-25 | 48 | Guyuan (Ningxia Province)  Sanmenxia (Henan Province)  Xining (Qinghai Province)  Jingdezhen (Jiangxi Province)  Huzhou (Zhejiang Province)  Longyan (Fujian Province)  Jiamusi and Mudanjiang (Heilongjiang Province)  Liaoyuan and Siping (Jilin Province)  Qinhuangdao and Rizhao (Shandong Province)  Chengde and Xingtai (Hebei Province)  Chuzhou and Huangshan (Anhui Province)  Guangyuan and Panzhihua (Sichuan Province)  Bijie and Tonghua (Guizhou Province)  Chaoyang, Dalian and Dandong (Liaoning Province)  Hohhot, Chifeng and Tongliao (Inner Mongolia)  Liuzhou, Qinzhou and Wuzhou (Guangxi Province)  Bazhong, Suining, Tongren and Yibin (Sichuan Province)  Baoshan, Lijiang, Yuxi and Zhaotong (Yunnan Province)  Taiyuan, Changzhi, Datong, Shuozhou, Yuncheng (Shanxi Province)  Qingyuan, Shantou, Shanwei, Shaoguan, Yangjiang and Zhanjiang (Guangdong Province) |

**Supporting Figure**

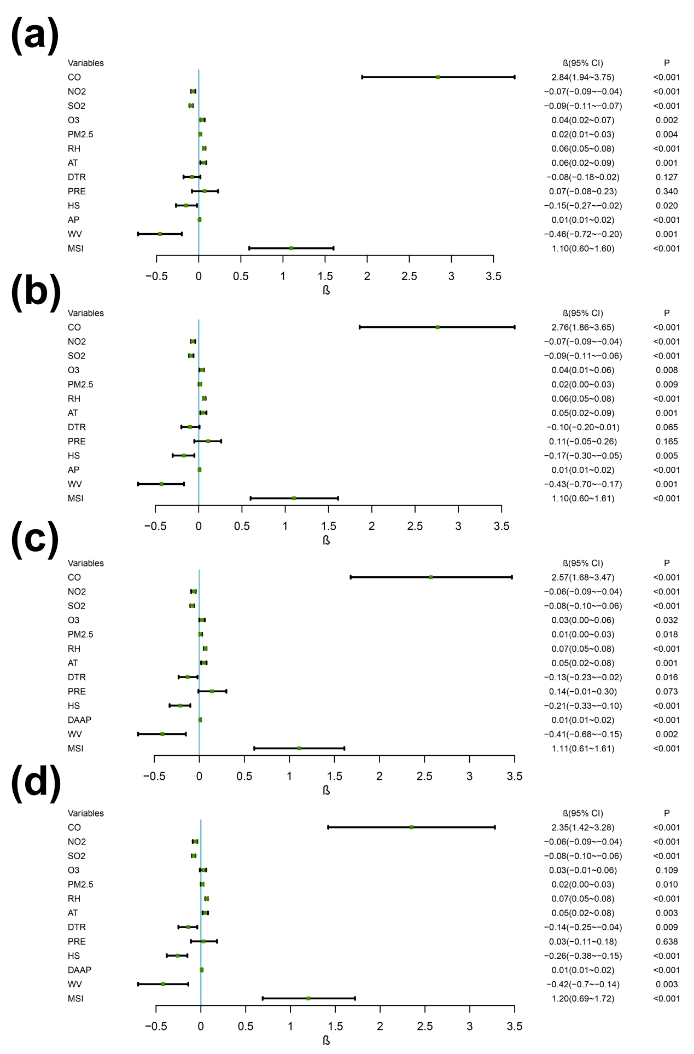

Figure S1 The relationship between air pollutants, meteorological factors and confirmed case counts in simple negative binomial regression model.
